# Seroprevalence of antibodies against SARS-CoV-2 virus in the adult Norwegian population, winter 2020/2021: pre-vaccination period

**DOI:** 10.1101/2021.03.23.21253730

**Authors:** Erik Eik Anda, Tonje Braaten, Kristin B. Borch, Therese H. Nøst, Sairah L. F. Chen, Marko Lukic, Eiliv Lund, Frode Forland, David Leon, Brita Askeland Winje, Anne-Marte Bakken Kran, Mette Kalager, Fridtjof Lund Johansen, Torkjel M. Sandanger

## Abstract

Since early 2020, over 123 million people worldwide have been diagnosed with coronavirus disease (Covid-19), but the true number of infections with severe acute respiratory syndrome coronavirus 2 (SARS-CoV-2) is undoubtedly higher. The seroprevalence of antibodies against SARS-CoV-2 can provide crucial epidemiological information about the extent of infections independent of virologically detected case numbers. There is no large population-based SARS-CoV-2 seroprevalence survey from Norway; thus we estimated SARS-CoV-2 seroprevalence in Norway before the introduction of vaccines and described its distribution across demographic groups. In November-December 2020, a total of 110,000 people aged 16 years or older were randomly selected from the National Population Register and invited to complete a questionnaire and provide a dried blood spot (DBS) sample. The response rate was 30%; compliance rate for return of DBS samples was 88%. The national weighted and adjusted seroprevalence was 0.9% (confidence interval 0.7-1.0).

Seroprevalence was highest among those aged 16-19 years (1.9%, 0.9-2.9), those born outside the Nordic countries 1.4% (1.0-1.9), and in the counties of Oslo 1.7 % (1.2-2.2) and Vestland 1.4% (0.9-1.8). The ratio of SARS-CoV-2 seroprevalence (0.9) to the cumulative incidence of virologically detected cases by mid-December 2020 (0.8%) was slightly above one. SARS-CoV-2 seroprevalence was low before the introduction of vaccines in Norway and was comparable to virologically detected cases, indicating that most cases in the first 10 months of the pandemic were detected. Preventive measures including contact tracing have been effective, people are complying with social distancing recommendations, and local efforts to contain outbreaks have been essential.

## Introduction

To-date an estimated 123.7 million people worldwide have been diagnosed with coronavirus disease (Covid-19) [1]. However, as these figures are based on the number of virologically detected cases of severe acute respiratory syndrome coronavirus 2 (SARS-CoV-2), they underestimate the true prevalence and incidence of Covid-19 due to limited test coverage, symptom-based test strategies, and the presence of asymptomatic cases [2, 3]. This underestimation limits our understanding of the burden of Covid-19 and impedes the development of effective public health strategies. The seroprevalence (i.e., the number of individuals with antibodies present in a defined population at a given time) of antibodies against SARS-CoV-2 can supply useful and needed estimates of the number of people that have been infected [4, 5]. Typically, antibodies appear in the blood within 4 weeks of detecting the presence of a microbe and serve as an indicator of past infection [6]. Although the level of SARS-CoV-2 antibodies is suspected to decline several months after infection [7], the window for antibody detection is far longer than that for viral detection.

A recent, large meta-analysis [8] reported varied SARS-CoV-2 seroprevalence, from 1.7% in the Western Pacific region, to 4.7% in the European region, to 19.6% in India. Moreover, the ratio of SARS-CoV-2 seroprevalence to the cumulative incidence of virologically detected cases was 8.4 in the European region, indicating that for each virologically detected SARS-CoV-2 case, at least eight remained undetected (Spearman’s rank correlation coefficient across all locations was 0.59) [8]. In Norway, 44 356 virologically detected SARS-CoV-2 cases had been reported by December 20, 2020, suggesting a cumulative incidence proportion of 0.8% [9].

To-date, no large study with a population-based random sample has estimated SARS-CoV-2 seroprevalence in Norway. Three smaller studies have estimated a seroprevalence of 1.0% and 0.6% in Norway, and 1.4% in Oslo [10, 11]. An accurate estimate of seroprevalence in Norway is important for ongoing Covid-19 containment and vaccination strategies, for estimating infection fatality rates, and for assessing the effectiveness of implemented restrictions or non-pharmaceutical interventions. Experiences from Norway’s low-density population setting may apply to other similar regions and could be valuable in creating strategies to manage Covid-19 going forward, and for future pandemics.

Thus, we aimed to estimate SARS-CoV-2 seroprevalence in a representative sample of inhabitants of Norway before the introduction of vaccines and to describe the distribution of this seroprevalence across relevant demographic groups.

## Material and methods

### Study population

This population-based, cross-sectional study included adults (over 16 years) in Norway. To be eligible, individuals had to have a national identity number, known country of birth, a registered address and a mobile phone number. Individuals living in prisons, nursing homes, or long-term psychiatric institutions (all of whom represent approximately 1% of the national population over 16 years of age [12]) were not eligible for inclusion. As previous population-based studies have demonstrated that response rates are not evenly distributed across age groups [13], we used a sampling frame from the Norwegian Institute of Public Health, which suggests oversampling of specific age groups, namely 16-19 years (x2), 20-29 years (x1.5), 65-74 years (x1.5), and 75+ years (x 2). Those born outside the Nordic countries were also oversampled (x2). Based on these methods, in November-December 2020, a total of 110 000 eligible individuals were randomly selected from the National Population Register and were sent an invitation via text message. 31 458 indicated their willingness to participate and were sent information about the study and were asked to complete an electronic or paper questionnaire and take a dried blood (DBS) spot sample. Of these, 27 700 completed the questionnaire and returned the DBS sample (Figure 1).

**Figure 1.**
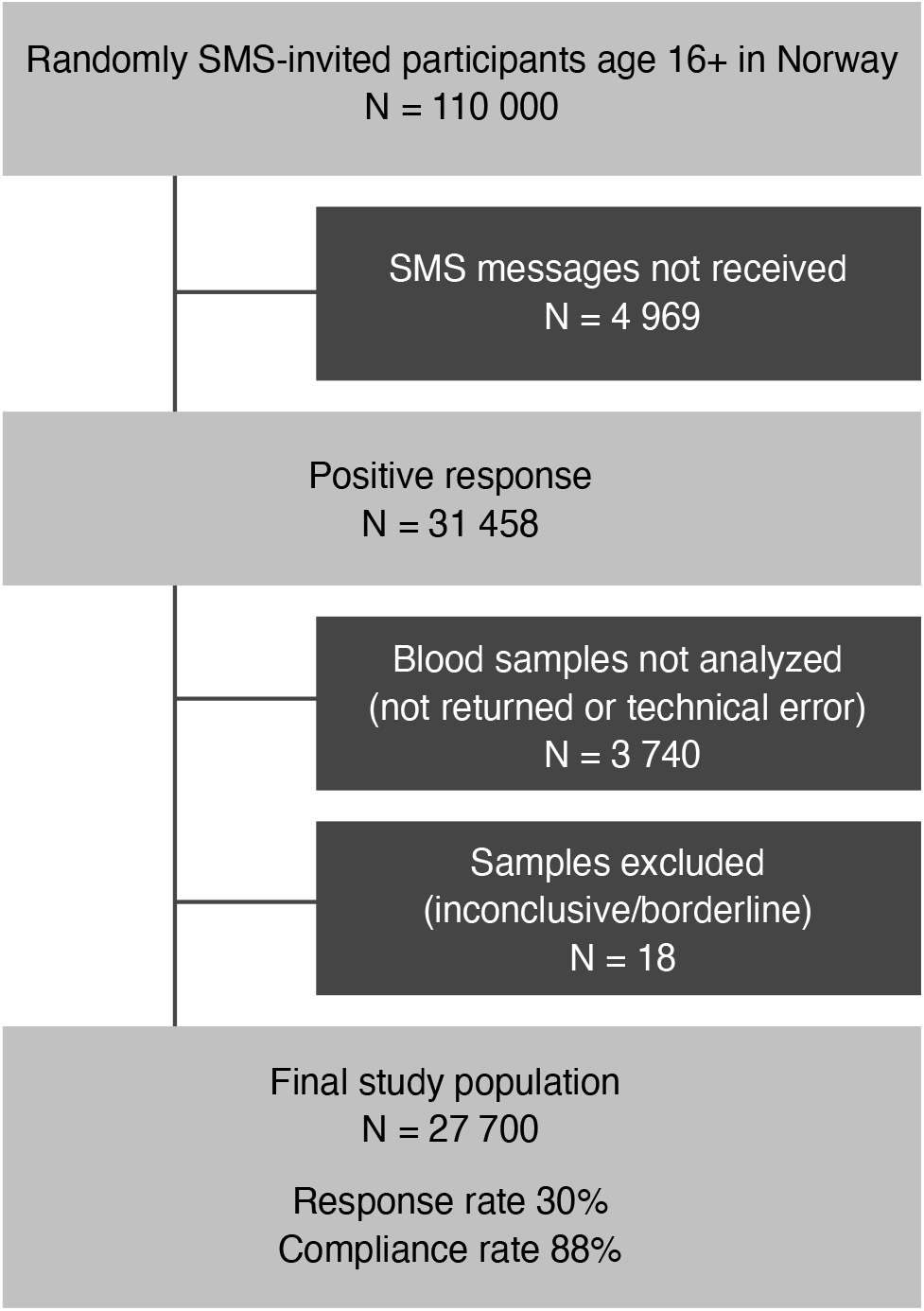
Flowchart of the invited population and final study sample.

## Ethical considerations

All participants gave written or electronic informed consent to participate in the study. The project group adheres to the Helsinki Declaration. This study has been approved by the Regional Committee for Medical Research Ethics, North Norway (reference number: 154985/2020) and the Norwegian Data Protection Authority (reference number: 758042/2020).

## Data collection

The questionnaire (see Supplements) collected information on education level, occupation, number of people living in the household, and Covid-19 infection, diagnosis, and symptoms. A total of 1505 participants did not complete the questionnaire. Information on age, sex, place of birth (Nordic countries or outside), and county of residence was extracted from Statistics Norway. Dried blood spot (DBS) samples were collected using a self-sampling kit for antibody testing (provided by VITAS - Analytical Services; a Good Manufacturing Practice-certified chemical analysis contract laboratory, Oslo, Norway). Information was provided on how to perform DBS sampling correctly, including online video instructions. Participants were to place captured capillary blood, after finger prick with a lancet, onto two circles on a filter paper card, which could collect 50 µL each. Participants then placed the filter paper card in a resealable bag containing desiccant, deposited that bag in a return envelope addressed to VITAS, and sent it by regular mail. DBS samples arrived between November 25, 2020 and February 15, 2021, with the bulk arriving in mid-to late-December. Upon receipt the samples were extracted at VITAS and then stored at −80°C until analysis.

## Detection of SARS-CoV-2 antibodies

Analyses of SARS-CoV-2 Immunoglobulin Gamma (IgG) antibodies were performed at VITAS, using an EUROIMMUN Anti-SARS-CoV-2 assay, an enzyme linked immunosorbent assay (ELISA) that provides semi-quantitative *in vitro* detection of human antibodies against the SARS-CoV-2 spike S1 protein. A test panel consisting of pre-pandemic samples and sera from PCR-confirmed Covid-19 convalescents showed >94% sensitivity and 99.9% specificity, but the sample size for that validation was limited (n=601). To minimize false positives, all samples that were positive or borderline by EUROIMMUN [14] underwent confirmatory analysis at the Department of Immunology at Oslo University Hospital. This consisted of a multiplexed bead-based flow cytometric assay and analyzed antibodies against receptor-binding domain (RBD) and the full-length spike protein. The cutoff was set to obtain a specificity of 99.9%; sensitivity was 84% and 92% when including borderline values. Analytical methods are described in more detail in the Supplementary Materials.

## Data treatment and statistical analyses

We defined SARS-CoV-2 IgG seropositivity as an EUROIMMUN value of >0.8, an RBD >5, and a Spike value >5, in addition to a signal for background noise/blank below 3000 in the multiplexed flow cytometric methods. However, if all three IgG thresholds were met, the sample was considered positive even if background noise slightly exceeded 3000. Twelve samples only contained enough blood for EUROIMMUN. Of these, four were positive, with values above 3.8, the highest observed among samples that were considered negative after flow cytometric analyses. We excluded samples with undetermined status (due to elevated blanks and below-threshold values in confirmatory analyses, or borderline EUROIMMUN results with too little blood for confirmatory analyses) (N=18 samples).

We dichotomized IgG antibodies (yes/no) in our statistical analysis. Seroprevalence was estimated for the total study sample, and by age group (16-19, 20-44, 45-66, 67-79, 80+ years), sex (women, men), place of birth (Nordic or outside Nordic countries), county (11 counties), educational level (primary school/junior high school, high school, vocational school, university or college), occupation (healthcare worker yes/no), and number of people living in the household (1, 2-4, 5-6). We evaluated the temporal stability of the seroprevalence estimate by dividing the sampling period into two shorter periods (before or after January 1, 2021). Overall and subgroup seroprevalence was calculated of the number of seropositive individuals divided by the number of individuals who returned DBS samples, and is presented as percentages with 95% confidence intervals (CI). We calculated the ratio of SARS-CoV-2 seroprevalence to the cumulative incidence of virologically detected cases registered at the Institute of Public Health [9] by December 20, 2020. We were not able to exclude data on individuals younger than 16 from virologically detected cases.

We used rake weighting to adjust population estimates of seroprevalence by age, sex, place of birth (Nordic or outside Nordic countries), and county based on individual-level data for the invited sample (participants and non-responders) together with the corresponding distributions from the source population, provided by the Norwegian Population Register. We applied propensity scores for nonresponse adjustment and jackknife replicate weights for the raking procedure [15]. The estimates were subsequently corrected for test performance [16]. We also retrieved population-level data on age, sex, place of birth, county of residence, education level, and occupation (healthcare worker yes/no) registered in Statistics Norway as of January 1, 2021. Correlation between weighted seroprevalence and cumulative incidence in the different counties at the time when most DBS samples were sent to the laboratory was calculated using Pearson’s correlation coefficient. All data treatment and statistical analyses were performed using STATA 16 [17].

## Results

Mean age in the study sample was 49.4 years. Participants aged 20-44 years comprised 36.8% of the sample, 40.5% were aged 45-66 years, 3.1% were aged younger than 20 years, and 2.8% were 80 years or older. More women (57.4%) than men (42.6%) participated, and 86.9% were born in Nordic countries. The distribution of participants by county was close to the national average. More than half of participants (52.5%) reported a university or college education level, and most (74.5%) had 2-4 people living in their household. Single-person households represented 16.7%, and those with five or more people represented 8.8%; 14.2% of participants reported that they were healthcare workers (Table 1).

**Table 1:** Descriptive summaries of the study sample, non-responders, and national numbers from Statistic Norway.

| Variables | Study sample<br>n=27 700 | Non-responders<br>n=76 937 | National, 16 years or<br>older* |
| --- | --- | --- | --- |
| Data from Statistics Norway |  |  |  |
| Age |  |  |  |
| Mean (SD) | 49.4 (17.3) | 47.5 (20.2) | NA |
| % (N) |  |  |  |
| 16-19 | 3.1 (868) | 6.3 (4834) | 5.7 |
| 20-44 | 36.8 (10 190) | 41.8 (32 186) | 40.7 |
| 45-66 | 40.5 (11 206) | 30.2 (23 202) | 34.3 |
| 67-79 | 16.8 (4655) | 14.6 (11 254) | 13.9 |
| 80+ | 2.8 (781) | 7.1 (5461) | 5.4 |
| Sex, % (N) |  |  |  |
| Women | 57.4 (15 895) | 47.8 (36 798) | 49.6 |
| Men | 42.6 (11 793) | 52.2 (40 134) | 50.4 |
| Place of birth, % (N) |  |  |  |
| Nordic countries | 86.9 (24 062) | 71.5 (54 976) | 82.5 |
| Outside Nordic countries | 13.1 (3626) | 28.5 (21 956) | 17.5 |
| Counties, % (N) |  |  | All (18+) |
| Troms/Finmark | 6.1 (1685) | 4.3 (3340) | 4.5 (4.6) |
| Nordland | 4.4 (1213) | 4.6 (3498) | 4.5 (4.5) |
| Trøndelag | 8.4 (2314) | 8.7 (6687) | 8.7 (8.8) |
| Møre og Romsdal | 4.5 (1256) | 5.1 (3908) | 4.9 (4.9) |
| Vestland | 12.0 (3326) | 11.4 (8777) | 11.8 (11.8) |
| Rogaland | 7.5 (2072) | 8.4 (6479) | 9.0 (8.7) |
| Agder | 4.9 (1362) | 5.9 (4524) | 5.7 (5.7) |
| Vestfold og Telemark | 7.8 (2155) | 8.0 (6126) | 7.8 (7.9) |
| Viken | 24.3 (6714) | 23.4 (17 977) | 23.2 (23.0) |
| Oslo | 13.7 (3782) | 13.1 (10 039) | 12.9 (13.2) |
| Innlandet | 6.5 (1806) | 7.3 (5577) | 6.9 (7.1) |
| Data from questionnaire |  |  |  |
| Education level, % (N) |  |  |  |
| Primary school/Junior high school | 7.7 (2022) | NA | 25.3 |
| High school | 25.9 (6776) | NA | 37.0 |
| Vocational school | 13.8 (3617) | NA | 3.0 |
| University or college | 52.5 (13 736) | NA | 34.6 |
| Healthcare worker, % (N) |  |  |  |
| Yes | 14.2 (3716) | NA | 22 |
| No | 85.8 (22 472) | NA | 78 |
| Number of people living in household,<br>N (%) |  |  |  |
| 1 | 16.7 (4376) | NA | 39.3 |
| 2-4 | 74.5 (19 500) | NA | 55.0 |
| 5+ | 8.8 (2312) | NA | 15.7 |

N-number, SD-standard deviation
\*Numbers from Statistics Norway [12]

By December 20, 2020, 44 356 individuals had been diagnosed with SARS-CoV-2 in Norway, for an estimated Covid-19 cumulative incidence proportion of 0.8 % [9] (Figure 2b). The weighted and adjusted SARS-CoV-2 seroprevalence was 0.9 % (CI 0.7-1.0) (Table 2). The crude and weighted estimates were similar in different subgroups, with the highest seroprevalence observed in those aged 16-19 years (1.9%, CI 0.9-2.9), among Norwegians born outside the Nordic countries 1.4% (CI 1.0-1.9), and in the counties of Oslo 1.7% (CI 1.2-2.2) and Vestland 1.4% (CI 0.9-1.8). The lowest seroprevalence was observed in the county of Møre and Romsdal 0% (CI 0.0 (0.0-0.4). Sex, education level, number of people living in the household, and occupation showed minor differences in seroprevalence between groups. The maps in Figures 2a and 2b show the spatial variation in weighted seroprevalence across the counties of Norway and the cumulative incidence of virologically confirmed cases in the same counties as of December 20, 2020. The ratio of seroprevalence to the cumulative incidence of virologically detected cases was 1.1 and the Pearson’s correlation coefficient between weighted seroprevalence and cumulative incidence per county was r=0.84. The estimated seroprevalence was 0.7% (0.6-0.9) before January 1, 2021 and 1.0% (0.9-1.2) after January 1, 2021.

**Table 2:** Seroprevalence in selected groups of the final study sample (November 2020-February 2021)

| <u>Variables</u> | <u>Positive tests</u><br><u>(N=)</u> | <u>Crude seroprevalence %</u><br><u>(95% CI)</u> | <u>Weighted seroprevalence</u><br><u>% (95% CI)</u> |
| --- | --- | --- | --- |
| **Data from Statistics Norway** |  |  |  |
| **Norway** | **234** | **0.8 (0.7-1.0)** | **0.9 (0.7-1.0)** |
| **Age** |  |  |  |
| **16-19** | 16 | 1.8 (1.1-3.0) | 1.9 (0.9-2.9) |
| **20-44** | 100 | 1.0 (0.8-1.2) | 1.0 (0.8-1.2) |
| **45-66** | 86 | 0.8 (0.6-1.0) | 0.7 (0.6-0.9) |
| **67-79** | 29 | 0.6 (0.4-0.9) | 0.5 (0.3-0.7) |
| **80+** | 3 | 0.4 (0.1-1.2) | 0.4 (0.0-0.9) |
| **Sex** |  |  |  |
| **Women** | 133 | 0.8 (0.7-1.0) | 0.8 (0.6-1.0) |
| **Men** | 101 | 0.9 (0.7-1.0) | 0.9 (0.7-1.1) |
| **Place of birth** |  |  |  |
| **Nordic countries** | 184 | 0.8 (0.7-0.9) | 0.7(0.6-0.9) |
| **Outside Nordic countries** | 50 | 1.4(1.0-1.8) | 1.4 (1.0-1.9) |
| **Counties** |  |  |  |
| **Troms/Finmark** | 12 | 0.7 (0.4-1.3) | 0.7 (0.2-1.1) |
| **Nordland** | 6 | 0.5 (0.3-1.1) | 0.4 (0.0-0.9) |
| **Trøndelag** | 8 | 0.4 (0.2-0.7) | 0.4 (0.0-0.8) |
| **Møre og Romsdal** | 1 | 0.1 (0.0-0.6) | 0.0 (0.0-0.1) |
| **Vestland** | 39 | 1.2 (0.9-1.6) | 1.4(0.9-1.8) |
| **Rogaland** | 15 | 0.7 (0.4-1.2) | 0.7 (0.3-1.2) |
| **Agder** | 9 | 0.7 (0.3-1.3) | 0.7 (0.1-1.2) |
| **Vestfold og Telemark** | 6 | 0.3 (0.1-0.6) | 0.2 (0.0-0.4) |
| **Viken** | 67 | 1.0 (0.8-1.3) | 1.0 (0.7-1.3) |
| **Oslo** | 60 | 1.6 (1.2-2.0) | 1.7(1.2-2.2) |
| **Innlandet** | 10 | 0.6 (0.3-1.0) | 0.6 (0.2-1.1) |
| **Data from questionnaire** |  |  |  |
| **Education level** |  |  |  |
| **Primary school/Junior high school** | 17 | 0.8 (0.5-1.3) |  |
| **High school** | 54 | 0.8 (0.6-1.0) |  |
| **Vocational school** | 22 | 0.6 (0.4-0.9) |  |
| **University or college** | 127 | 0.9 (0.8-1.1) |  |
| **Healthcare worker** |  |  |  |
| **Yes** | 42 | 1.1 (0.8-1.5) |  |
| **No** | 178 | 0.8 (0.7-0.9) |  |
| **Number of people living in household** |  |  |  |
| **1** | 37 | 0.8 (0.6-1.2) |  |
| **2-4** | 161 | 0.8 (0.7-1.0) |  |
| **5+** | 22 | 1.0 (0.6-1.4) |  |

CI- confidence interval, N- number

**Figure 2.**
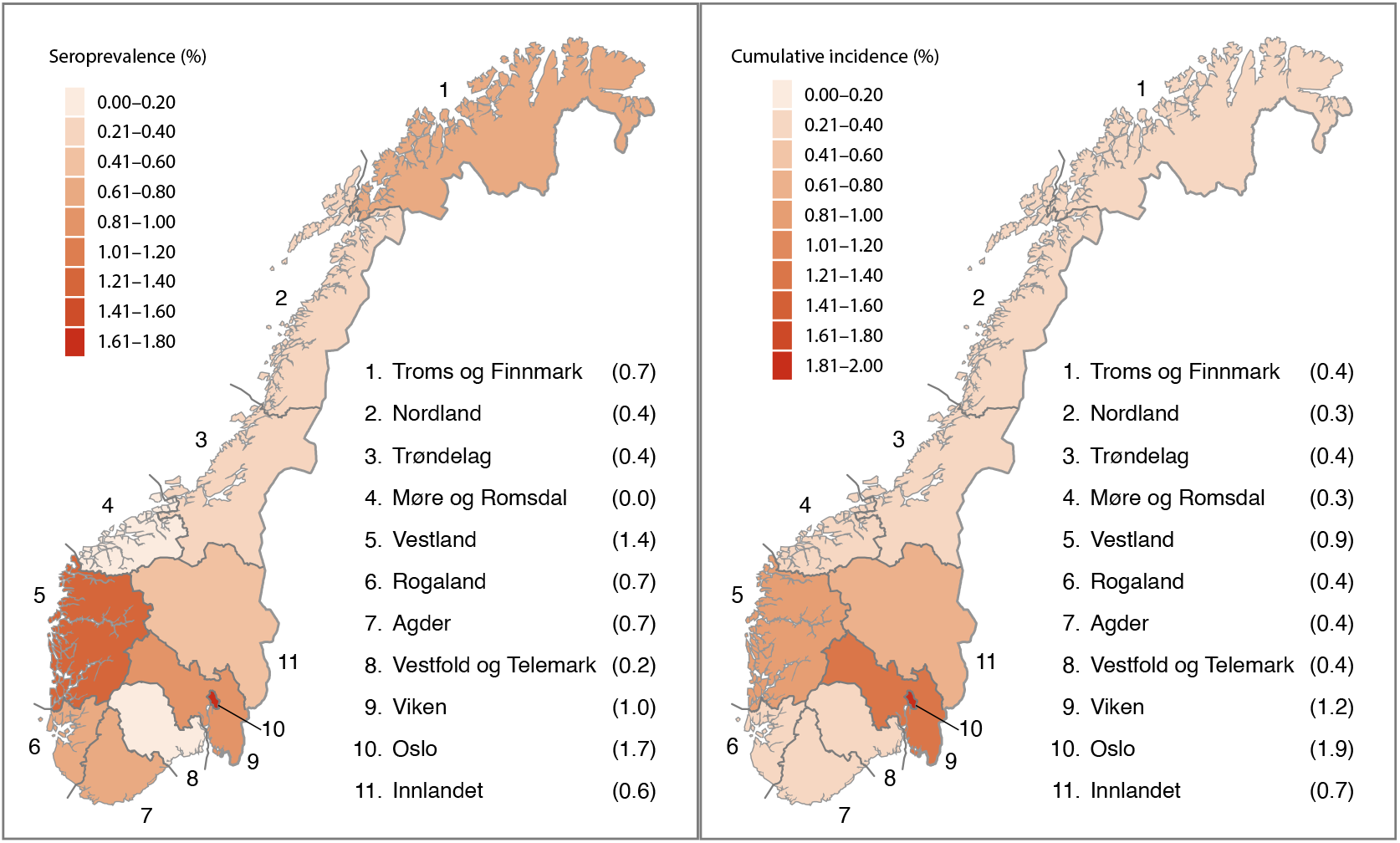
A) Weighted seroprevalence (%) and B) cumulative incidence of SARS-CoV-2 virus in Norway by county. The median sampling period was late December 2020 and cumulative incidence was obtained for December 20, 2020. Map provided by the Norwegian Mapping Authority, modified by UiT The Arctic University of Norway.

## Discussion

### Summary of main findings

In this random-sample, population-based study, we found a low SARS-CoV-2 seroprevalence (0.9%) in the Norwegian population by January 2021. Seroprevalence was highest in those aged 16-19 years (1.9%), in Oslo County (1.7%), and among persons born outside the Nordic countries (1.4%). There were considerable geographical differences, with the lowest seroprevalence observed in Møre and Romsdal County (0%). Finally, we found a low ratio (1.1) of seroprevalence to the cumulative incidence of virologically detected cases, indicating that a substantial number of the Covid-19 cases in Norway are detected [8].

### National seroprevalence in the context of other Norwegian studies

Our observed national seroprevalence (0.9%) is supported by two studies performed in residual clinical samples from hospital laboratories in Norway, which reported national seroprevalences of 1.0% in spring [11] and 1.7% in and early autumn [10] of 2020. We saw substantial geographical differences, with the highest seroprevalences observed in counties with the two largest cities in Norway – Oslo and Bergen, and previous seroprevalence studies from Oslo support our results [18]. The airborne and close-contact nature of Covid-19 transmission; factors related to high population density in urban settings, such as frequent human interactions and a high number of national and international flights; and younger population distributions, may explain these results. Moreover, the key public health strategy of test, trace, and isolate is more challenging in large cities than in smaller communities.

Seroprevalence was highest in the youngest age group, a trend that was also found in the UK, where the highest seroprevalence was reported in those aged 18-24 years (7.9%) [19]. We speculate that young adults have more frequent contact with other individuals, and in larger groups, thus facilitating infection. This contact could be attributable to school, greater social needs, or the higher concentrations of young people living in Norway’s urban centers, which likely increases the risk of Covid-19 infection regardless of age. On average, young adults infected with Covid-19 experience milder symptoms than older adults [20], which may result in a lower motivation to be tested and/or to reduce social contact.

We observed a higher seroprevalence among persons born outside Nordic countries (1.4 %). This group is over-represented in Norway’s larger cities, indicating a possible contributing role of population density in driving higher seroprevalence. However, we may not have achieved national representativeness for this group, as the proportion of persons in our study sample with higher education and born outside of the Nordic countries was greater than in the general population [12]. Language barriers and skepticism towards the SMS invitation are plausible reasons for not capturing a representative sample of this group.

In contrast to the higher seroprevalence observed among healthcare workers in many other countries [8], seroprevalences were similar in healthcare and non-healthcare workers in our study. Low infection rates, few hospitalized patients, and good access to personal protective equipment likely explain our result. There may be a subtle difference between the seroprevalences of healthcare workers in high-risk settings compared to other healthcare workers, but our study did not differentiate between these groups.

Differences in education level did not appear to affect seroprevalence. This variable is highly age-dependent, as lower age usually implies lower completed education level. Despite this, our lower education group did not have higher seroprevalence. Lastly, in contrast to earlier findings [19], we found minor differences in seroprevalence between those living alone and those living in larger households.

### Comparisons to other countries

When comparing the Norwegian seroprevalence of 0.9% to those of the serological studies assessed in the recent review by [8], it is evident that Norway has one of the lowest seroprevalences globally. The estimated pooled seroprevalence in the world general population as of December 22, 2020 was 8.0% (95% CI 6.8-9.2), with the lowest observed in the Western Pacific region (1.7%, 95% CI 0.0-5.0), and an average of 4.7% (95% CI 3.6-5.9) in the European region [8]. In our study and in a UK sample, the highest seroprevalence was found in the youngest age group [19]. However, in the review by Chen et al [8], this age group was reported to have the lowest seroprevalence globally (2.1%). Whether this is a coincidence or reflects similar infection rates globally in this age group is unknown.

Both the correlation (r=0.84) between and the ratio (1.1) of seroprevalence to the cumulative incidence confirm the low incidence of SARS-Cov-2 in Norway. Globally, none of the countries included in the meta-analysis had a ratio lower than or equal to 1 and the European average was 8.4. The lowest ratio was found in the state of Utah in the US (2.4), and the highest ratio was found in India (56.5) [8], likely due to variations in testing capacities and opportunities in the different countries. In addition, the regression coefficient describing the relationship between seroprevalence and reported cases stratified by county is high enough to indicate low numbers of undetected cases in Norway. Possible explanations for the low seroprevalence in Norway are low population density in most parts of the country, adherence to national and local restrictions, effective testing, contact tracing, and isolation strategies, and the mobilization of financial and systematic resources.

### Strengths and limitations

The strengths of our study include the random sample, population-based study design, high completion rate of DBS samples (88%), and the low risk of overestimation of seroprevalence. There are no stability issues related to the time elapsed from DBS sampling to time of arrival at the laboratory. However, this study also has limitations. The response rate was 30%; but the age and sex distributions were similar to that of the Norwegian general population, so the overall results may still be representative of the population. However, even though we oversampled, the results from some subgroups may be more uncertain and not representative of the Norwegian population.

Studies involving personal information linked to risk behavior tend to have a lower response from high-risk groups [21]. However, having antibodies is not necessarily linked to a stigma or viewed as high risk. Because of the low response rate in the youngest age group and because this group had the highest seroprevalence, the estimates could be too low. On the other hand, weighting and adjustments for sensitivity and specificity of results reduced underreporting to a minimum.

There are several reasons why seroprevalence may be underestimated in our study. First, those who were very recently infected, were less likely to have detectable antibody levels at the time of sampling [22]. Second, because of the sensitivity of the two tests (94% and 84%), some false negative results were expected, but due to the low prevalence in general, these cases are few.

Furthermore, the properties of the tests used to detect antibodies might differ between populations with a different case-mix [23]. It is possible that in a region/population in which the proportion of asymptomatic cases was high, the tests would perform with somewhat lower sensitivity, resulting in an underestimated seroprevalence in that specific population. The proportion of participants with only Primary school/Junior high school was low in our study population. Lastly, we did not tested children under the age of 16 years, and we do not know how the addition of these children would have affected the estimated seroprevalence, but this is only relevant when comparing seroprevalence with cumulative incidence, not when estimating seroprevalence in the adult population.

## Conclusion

Although there are limitations to seroprevalence estimates, such as time between infection and antibody testing (waning antibodies over time or testing before antibodies develop) and individual antibody response to the infection, IgG antibody seroprevalence is probably the best indication of population protection and past infection irrespective of SARS-CoV-2 test capacity or availability in the population.

SARS-CoV-2 seroprevalence in Norway was low during winter 2020/2021, before vaccines were introduced. Our findings suggest that the proportion of virologically undetected cases is low. The low seroprevalence leaves Norway particularly vulnerable to a third wave during the Spring of 2021, as there is no indication of past infection on a population level that might confer protection. However, the progress of the ongoing vaccination campaign will rectify this, and a repeat seroprevalence survey should be conducted later in 2021.

## Supporting information

Questionnaire

Analytical methods

## Data Availability

The datasets analysed during the current study are not publicly available due to local and national ethical and security policies, but can be obtained upon request from the corresponding author in an anonymized format after the study has been published.

## Acknowledgements

The authors would like to thank the participants for their valuable contributions to this survey. We acknowledge the valuable and enthusiastic efforts provided by Bente A. Augdal, Merete Albertsen, Karina Standahl Olsen, Hasse Melbye, Paolo Vineis, Filippo Bianchi, and involved colleagues at our department. We also thank VITAS for their services.

## Author contributions

T.M.S., E.E.A., T.B., K.B.B., T.H.N. and S.L.F.C. conceived of the presented idea, planned the sample collections and have written the manuscript. M.L. and T.B. conducted the statistical analyses. All authors helped shape the research, analysis, discussed the results and contributed to the final manuscript.

## References

1. Johns Hopkins University and Medicine [Internet]. COVID-19 Dashboard by the Center for Systems Science and Engineering (CSSE) at Johns Hopkins University (JHU) [Cited 21.03.23]. Available from: https://coronavirus.jhu.edu/map.html.

2. Eckerle I, Meyer B. SARS-CoV-2 seroprevalence in COVID-19 hotspots. The Lancet. 2020;396(10250):514–5.

3. Oran DP, Topol EJ. The Proportion of SARS-CoV-2 Infections That Are Asymptomatic : A Systematic Review. Ann Intern Med. 2021.

4. Lai CC, Wang JH, Hsueh PR. Population-based seroprevalence surveys of anti-SARS-CoV-2 antibody: An up-to-date review. Int J Infect Dis. 2020;101:314–22.

5. Jacofsky D, Jacofsky EM, Jacofsky M. Understanding Antibody Testing for COVID-19. The Journal of Arthroplasty. 2020;35(7):S74–S81.

6. Murin CD, Wilson IA, Ward AB. Antibody responses to viral infections: a structural perspective across three different enveloped viruses. Nat Microbiol. 2019;4(5):734–47.

7. Wang Y, Li J, Li H, Lei P, Shen G, Yang C. Persistence of SARS-CoV-2-specific antibodies in COVID-19 patients. Int Immunopharmacol. 2021;90:107271.

8. Chen X, Chen Z, Azman AS, Deng X, Sun R, Zhao Z, et al. Serological evidence of human infection with SARS-CoV-2: a systematic review and meta-analysis. The Lancet Global Health. 2021.

9. Norwegian Institute of Public Health [Internet]. Statistikk om koronavirus og covid-19 (Statistics about coronavirus and Covid-19) 2020 [Cited 21.03.04]. Available from: https://www.fhi.no/sv/smittsomme-sykdommer/corona/dags--og-ukerapporter/dags--og-ukerapporter-om-koronavirus/.

10. Tunheim G, Bakken A-M, Rø KG, Hungnes O, Lund-Johansen F, Tran T, et al. Seroprevalence of SARS-CoV-2 in the Norwegian population measured in residual sera collected in late summer 2020. Oslo Norwegian Institute for Public Health; 2020.

11. Tunheim G, Kran A-MB, Rø G, Steens A, Hungnes O, Lund-Johansen F, et al. Seroprevalence of SARS-CoV-2 in the Norwegian population measured in residual sera collected in April/May 2020 and August 2019. Oslo; 2020

12. Statistics Norway [Internet]. Befolkningen (Population) 2021 [Cited 21.03.22]. Available from: https://www.ssb.no/befolkning/faktaside/befolkningen.

13. Abrahamsen R, Svendsen MV, Henneberger PK, Gundersen GF, Toren K, Kongerud J, et al. Non-response in a cross-sectional study of respiratory health in Norway. BMJ Open. 2016;6(1):e009912.

14. Zava TT, Zava DT. Validation of dried blood spot sample modifications to two commercially available COVID-19 IgG antibody immunoassays. Bioanalysis. 2020;13(1):13–28.

15. Valliant R, Dever JA. Survey Weights: A Step-by-step Guide to Calculation: Stata Press; 2018.

16. Diggle PJ. Estimating Prevalence Using an Imperfect Test. Epidemiology Research International. 2011;2011:1–5.

17. StataCorp. Stata Statistical Software: Release 16. College Station, TX: StataCorp LLC; 2019.

18. Helsingen LM, Løberg M, Refsum E, Gjøstein DK, Wieszczy P, Olsvik Ø, et al. A Randomised Trial of Covid-19 Transmission in Training Facilities. medRxiv 2020.

19. Ward H, Atchison C, Whitaker M, Ainslie KEC, Elliott J, Okell L, et al. SARS-CoV-2 antibody prevalence in England following the first peak of the pandemic. Nat Commun. 2021;12(1):905.

20. European Centre for Disease Prevention and Control. COVID-19 in children and the role of school settings in transmission - first update Stockholm: ECDC; 2020 [

21. Cheung KL, Ten Klooster PM, Smit C, de Vries H, Pieterse ME. The impact of non-response bias due to sampling in public health studies: A comparison of voluntary versus mandatory recruitment in a Dutch national survey on adolescent health. BMC Public Health. 2017;17(1):276.

22. Hansen CH, Michlmayr D, Gubbels SM, Mølbak K, Ethelberg S. Assessment of protection against reinfection with SARS-CoV-2 among 4 million PCR-tested individuals in Denmark in 2020: a population-level observational study. The Lancet. 2021.

23. Willis BH. Spectrum bias—why clinicians need to be cautious when applying diagnostic test studies. Family Practice. 2008;25(5):390–6.

