## Supplementary material for "Seroprevalence of antibodies against SARS-CoV-2 virus in the adult Norwegian population, winter 2020/2021: pre-vaccination period": Questionnaire

#### COVID-19 and Immunity in Norway

**Spørreskjema for sensitive data (TSD)**

Skjema er koblet til TSD (p1324)

**Sist endret**

2. desember 2020
23:16
av Tonje Braaten



### COVID-19 and Immunity in Norway

Side 1

#### COVID-19 AND IMMUNITY IN NORWAY (Korona og immunitet i Norge). A study on blood antibodies and immunity in the Norwegian populatio

Sideskift

Side 2

Mandatory questions are marked with *

#### Covid-19

Have you been tested for Covid-19? *

- Yes, at least one test was positive
- Yes, but the test was negative
- Yes, waiting for result
- No, I have not been tested

#### When was the first time you tested positive? *

Dette elementet vises kun dersom alternativet «Yes, at least one test was positive» er valgt i spørsmålet «Have you been tested for Covid-19?»

- February-March
- April-May
- June-July
- August-September
- October-November

#### Do you think you could have been infected with Covid-19? *

Dette elementet vises kun dersom alternativet «Yes, but the test was negative», «No, I have not been tested» eller «Yes, waiting for result» er valgt i spørsmålet «Have you been tested for Covid-19?»

- No
- Yes
- Do not know / do not wish to answer

#### Did you have Covid-19 symptoms when you were infected? *

Dette elementet vises kun dersom alternativet «Yes, at least one test was positive» er valgt i spørsmålet «Have you been tested for Covid-19?»

No

Yes

Do not know / do not wish to answer

#### Which symptoms did you have? *

Dette elementet vises kun dersom alternativet «Yes» er valgt i spørsmålet «Did you have Covid-19 symptoms when you were infected?»

- Cold/flu symptoms such as coughing or runny nose
- Difficulty breathing (more than usual)
- Fever
- Muscle or joint pain
- Loss of sense of smell or taste
- Stomach pain, nausea, or diarrhea
- Other symptoms

#### In which month did you experience the symptoms? *

Dette elementet vises kun dersom alternativet «Yes» er valgt i spørsmålet «Did you have Covid-19 symptoms when you were infected?»

Velg…

#### How sick were you? *

Dette elementet vises kun dersom alternativet «Yes» er valgt i spørsmålet «Did you have Covid-19 symptoms when you were infected?»

- Mildly sick
- Moderately sick
- Very sick
- Do not know / do not wish to answer

#### Were you hospitalised? *

Dette elementet vises kun dersom alternativet «Yes» er valgt i spørsmålet «Did you have Covid-19 symptoms when you were infected?»

- No
- Yes

#### Have you had symptoms that could be due to Covid-19 during 2020? *

Dette elementet vises kun dersom alternativet «Yes, but the test was negative», «No, I have not been tested» eller «Yes, waiting for result» er valgt i spørsmålet «Have you been tested for Covid-19?»

- No
- Yes
- Do not know / do not wish to answer

#### Which symptoms did you have? *

Dette elementet vises kun dersom alternativet «Yes» er valgt i spørsmålet «Have you had symptoms that could be due to Covid-19 during 2020?»

- Cold/flu symptoms such as coughing or runny nose
- Difficulty breathing (more than usual)
- Fever
- Muscle or joint pain
- Loss of sense of smell or taste
- Stomach pain, nausea, or diarrhea
- Other symptoms

#### In which month did you experience the symptoms? *

Dette elementet vises kun dersom alternativet «Yes» er valgt i spørsmålet «Have you had symptoms that could be due to Covid-19 during 2020?»

Velg…

#### How sick were you? *

Dette elementet vises kun dersom alternativet «Yes» er valgt i spørsmålet «Have you had symptoms that could be due to Covid-19 during 2020?»

- Mildly sick

Moderately sick

Very sick

Do not know / do not wish to answer

#### Were you hospitalised? *

Dette elementet vises kun dersom alternativet «Yes» er valgt i spørsmålet «Have you had symptoms that could be due to Covid-19 during 2020?»

No

Yes

Sideskift

Side 3

#### Do you currently have any of the following diseases/disorders? *

Heart disease

Cancer

Asthma

Chronic obstructive pulmonary disease (COPD) or other chronic lung disease

Liver disease

Kidney disease

- [ ] Central nervous system disorder
- [ ] Eating disorder
- [ ] High blood pressure
- [ ] Diabetes
- [ ] Rheumatic disorder
- [ ] Other autoimmune disease
- [ ] I do not have any of these diseases/disorders
- [ ] Do not know / do not wish to answer

Which type of diabetes do you have? *

Dette elementet vises kun dersom alternativet «Diabetes» er valgt i spørsmålet «Do you currently have any of the following diseases/disorders?»

- Type 1
- Type 2
- Gestational diabetes

Sideskift

Side 4

### Travel

How many days per week did you usually take public transport between January and March

2020? *

0

1

2 or more

How many days per week have you usually taken public transport since August 2020? *

0

1

2 or more

Sideskift

Side 5

Did you travel/were you traveling outside of Norway in February or March 2020? *

No

Yes

Which continent(s) did you travel to? *

Dette elementet vises kun dersom alternativet «Yes» er valgt i spørsmålet «Did you travel/were you traveling outside of Norway in February or March 2020?»

Europe

Asia

Africa

America

Oceania

#### Which country/countries in Europe did you travel to? *

Dette elementet vises kun dersom alternativet «Europe» er valgt i spørsmålet «Which continent(s) did you travel to?»

Sweden

Denmark

Finland

Great Britain

Germany

Austria

Italy

Spain

France

Poland

Lithuania

Other country in Europe

Were you in quarantine upon arriving back in Norway from your travel abroad in February or March 2020? *

?

Dette elementet vises kun dersom alternativet «Yes» er valgt i spørsmålet «Did you travel/were you traveling outside of Norway in February or March 2020?»

No

Yes

Have you traveled outside of Norway after March 2020? *

No

Yes

Which continent(s) did you travel to? *

?

Dette elementet vises kun dersom alternativet «Yes» er valgt i spørsmålet «Have you traveled outside of Norway after March 2020?»

Europe

Asia

Africa

America

Oceania

#### Which country/countries in Europe did you travel to? *

Dette elementet vises kun dersom alternativet «Europe» er valgt i spørsmålet «Which continent(s) did you travel to?»

- [ ] Sweden
- [ ] Denmark
- [ ] Finland
- [ ] Great Britain
- [ ] Germany
- [ ] Austria
- [ ] Italy
- [ ] Spain
- [ ] France
- [ ] Poland
- [ ] Lithuania
- [ ] Other country in Europe

#### Were you in quarantine upon arriving back in Norway from your travel abroad since March 2020? *

Dette elementet vises kun dersom alternativet «Yes» er valgt i spørsmålet «Have you traveled outside of Norway after March 2020?»

- No
- Yes

Sideskift

Side 6

Did you visit counties in Norway other than your home-county in February or March 2020? *

- No
- Yes

Which county/counties did you visit? *

Dette elementet vises kun dersom alternativet «Yes» er valgt i spørsmålet «Did you visit counties in Norway other than your home-county in February or March 2020?»

- Agder
- Innlandet
- Møre and Romsdal
- Nordland
- Oslo

Rogaland

Vestfold and Telemark

Troms and Finnmark

Trøndelag

Vestland (except Bergen)

Bergen

Viken

#### Did you visit counties in Norway other than your home-county after March 2020? *

No

Yes

#### Which county/counties did you visit? *

Dette elementet vises kun dersom alternativet «Yes» er valgt i spørsmålet «Did you visit counties in Norway other than your home-county after March 2020?»

Agder

Innlandet

Møre and Romsdal

Nordland

Oslo

Rogaland

Vestfold and Telemark

Troms and Finnmark

Trøndelag

Vestland (except Bergen)

Bergen

Viken

Sideskift

Side 7

### Education and employment

What is your highest completed level of education? *

Primary school / Junior high school

High school

Vocational school

University or college

Are you a student? *

No

Yes

How many days per week were you usually at school/college/university between March and May 2020? *

Dette elementet vises kun dersom alternativet «Yes» er valgt i spørsmålet «Are you a student?»

0

1

2 or more

How many days per week have you usually been at school/college/university since August 2020? *

Dette elementet vises kun dersom alternativet «Yes» er valgt i spørsmålet «Are you a student?»

0

1

2 or more

Are you employed? *

Yes

I am laid off

- Yes, but I am on sick leave
- I am retired
- No

How many days per week were you usually at your workplace (outside of the home) between March and May 2020? *

Dette elementet vises kun dersom alternativet «Yes», «Yes, but I am on sick leave» eller «I am laid off» er valgt i spørsmålet «Are you employed?»

- 0
- 1
- 2 or more

How many days per week have you usually been at your workplace (outside of the home) since August 2020? *

Dette elementet vises kun dersom alternativet «Yes», «Yes, but I am on sick leave» eller «I am laid off» er valgt i spørsmålet «Are you employed?»

- 0
- 1
- 2 eller more

In which sector/industry do you work? *

Dette elementet vises kun dersom alternativet «Yes», «Yes, but I am on sick leave» eller «I am laid off» er valgt i spørsmålet «Are you employed?»

- Healthcare
- Passenger transportation
- Sales and service
- Manufacturing
- Agriculture and fishing
- First response (fire, rescue, or police)
- Kindergarten and primary school
- Junior high school, high school, and higher education
- Other sector/industry

#### Do you work in *

Dette elementet vises kun dersom alternativet «Healthcare» er valgt i spørsmålet «In which sector/industry do you work?»

- Municipal healthcare
- Specialist healthcare
- Private healthcare

Sideskift

### Living conditions

How many people live in your household? *

- 1
- 2
- 3
- 4
- 5
- 6 or more

How many children (0-15 years) do you live with? *

?

Dette elementet vises kun dersom alternativet «2», «4», «3», «6 or more» eller «5» er valgt i spørsmålet «How many people live in your household?»

- 0
- 1
- 2
- 3
- 4 or more

Do you live with a child who goes to Kindergarten? *

Dette elementet vises kun dersom alternativet «3», «2», «4 or more» eller «1» er valgt i spørsmålet «How many children (0-15 years) do you live with?»

- No
- Yes

Do you live with a child who goes to school? *

Dette elementet vises kun dersom alternativet «3», «2», «4 or more» eller «1» er valgt i spørsmålet «How many children (0-15 years) do you live with?»

- No
- Yes

Do you live in an apartment complex with a shared entrance? *

- No
- Yes

Has anyone else in your household been tested positive for Covid-19? *

- No
- Yes
- Don't know / do not wish to answer

Has anyone in your social circle (friends, colleagues) been tested positive for Covid-19? *

- No
- Yes
- Don't know / do not wish to answer

Sideskift

Side 9

#### Other questions

How tall are you (in cm)? *

How much do you weigh (in kg)? *

Do you or have you previously smoked daily? *

- No
- Previously smoked
- Currently smoke
- Do not wish to answer

How many years ago did you quit smoking? *

Dette elementet vises kun dersom alternativet «Previously smoked» er valgt i spørsmålet «Do you or have you previously smoked daily?»

- 0-1
- 1-5
- 5-10
- More than 10
- Do not wish to answer

#### Do you use snuff? *

- No
- Yes
- Do not wish to answer

#### How often do you use snuff? *

Dette elementet vises kun dersom alternativet «Yes» er valgt i spørsmålet «Do you use snuff?»

- Everday
- A few times per week
- A few times per month
- Rarely

Have you used antibiotics in 2020? *

No

Yes

Do not know / do not wish to answer

Have you used steroids (e.g. Prednisolon) in 2020? *

No

Yes

Do not know / do not wish to answer

Did you get vaccinated for the current flu/influenza season (autumn 2020/2021)? *

No

Yes

Do you take cod liver oil or vitamin D supplements? *

No

Yes

Occasionally

Are you physically active at a moderate intensity level (slightly out-of-breath) at least 2.5 hours per week? *

No

Yes

Do not know / do not wish to answer

Do you consent to being contacted again for future studies on Covid-19? *

Yes

No

Se nylige endringer i Nettskjema

**Vilkår**
Personvern og vilkår for bruk
Nettskjema bruker informasjonskapsler
Tilgjengelighetserklæring

**Kontaktinformasjon**
Kontaktpunkter Nettskjema

**Ansvarlig for denne tjenesten**
Webseksjonen – USIT
