## Supplementary material for "Seroprevalence of antibodies against SARS-CoV-2 virus in the adult Norwegian population, winter 2020/2021: pre-vaccination period": Analytical methods

This supplementary material is hosted by *Eurosurveillance* as supporting information alongside the article **Seroprevalence of antibodies against SARS-CoV-2 virus in the adult Norwegian population, winter 2020/2021: pre-vaccination period**, on behalf of the authors, who remain responsible for the accuracy and appropriateness of the content. The same standards for ethics, copyright, attributions and permissions as for the article apply. Supplements are not edited by *Eurosurveillance* and the journal is not responsible for the maintenance of any links or email addresses provided therein.

### Analytical methods for detection of SARS-CoV2 antibodies

Antibody measurement: A multiplexed bead-based flow cytometric assay, referred to as microsphere affinity proteomics (MAP), was adapted for detection of SARS-CoV2 antibodies [1]. Thus amine-functionalized polymer beads were color-coded with fluorescent dyes as described earlier and reacted successively with amine-reactive biotin (sulfo-NHS-LC-biotin, Proteochem, USA) and neutravidin (Thermo Fisher). A DNA construct encoding the receptor-binding domain of Spike-1 protein (RBD) from SARS-CoV2 was provided by Florian Krammer, and the protocol described in [2] was used to produce recombinant protein in Expi293F cells [2]. Bacterially expressed full length nucleocapsid from SARS-CoV2 was purchased from Prospec Bio (www.prospecbio.com). Viral proteins solubilized in PBS were biotinylated chemically using a four to one molar ratio of sulfo-NHS-LC-biotin to protein. Free biotin was removed with G50 sephadex spin columns. Biotinylated proteins were bound to neutravidin-coupled microspheres with fluorescent barcodes. Beads with Neutravidin only were used as reference for background binding. Eluates from dried blood spots were incubated with a mixture of antigen-coupled and Neutravidin-only beads for 1h at 22ºC under constant agitation. The beads were washed twice in PBT, labelled with R-Phycoerythrin-conjugated goat-anti-Human IgG-Fc (Jackson Immunoresearch) for 20 min, washed again and analyzed by flow cytometry (Attune Next, Thermo Fisher). Specific binding was measured as the ratio of R-Phycoerythrin fluorescence intensity of antigen-coupled beads and neutravidin-only beads. Samples containing antibodies both to Nucleocapsid and RBD were considered to be positive. Reference panels containing samples from 287 individuals with PCR-confirmed SARS-CoV2 infection and 1343 pre-pandemic samples were used to set the cutoff. With a cutoff set to obtain a specificity of 100%, the sensitivity was 84% and 92% when including borderline values. The cutoff values for positives were five and ten for antibodies to RBD and Nucleocapsid, respectively. Borderline was defined as RBD higher than five.

**References**

1. Wu, W., Slastad, H., de la Rosa Carrillo, D., Frey, T., Tjonnfjord, G., Boretti, E., Aasheim, H.C., Horejsi, V. & Lund-Johansen, F. Antibody array analysis with label-based detection and resolution of protein size. *Mol Cell Proteomics* **8**, 245-257 (2009).

2. Amanat, F., Stadlbauer, D., Strohmeier, S., Nguyen, T.H., Chromikova, V., McMahon, M., Jiang, K., Arunkumar, G.A., Jurczyszak, D. & Polanco, J. A serological assay to detect SARS-CoV-2 seroconversion in humans. *Nature medicine*, 1-4 (2020).
